# The impact of prenatal alcohol exposure on sleep outcomes in 10,336 young adolescents: An Adolescent Brain Cognitive Development (ABCD) Study

**DOI:** 10.1101/2025.05.14.25327575

**Authors:** Emma K Devine, ReJoyce Green, Rachel Visontay, Hollie Byrne, Julia Riches, Elizabeth J. Elliott, Nicola C. Newton, Lindsay M. Squeglia, Louise Mewton, Lexine A. Stapinski

## Abstract

**Study Objectives:** This study investigated the associations between prenatal alcohol exposure (PAE), including low and moderate levels of exposure, and sleep outcomes in adolescence. This is an area that remains understudied despite evidence linking PAE to poor sleep in younger children and the growing recognition of harms associated with low levels of PAE.

**Methods:** Participants were 10,336 adolescents (aged 12-13) from the fourth assessment wave of the Adolescent Brain Cognitive Development Study. Cross-sectional generalised linear mixed models and generalised additive mixed models were used to assess the impact of prenatal alcohol exposure, conceptualised as the presence and absence of PAE, total drinks consumed during pregnancy (i.e. dose), and patterns of PAE (i.e., abstainers, light reducing, light stable, heavy reducing), on adolescent sleep outcomes.

**Results:** Adolescents with any PAE experienced worse sleep outcomes compared to those without, with the sleep-wake transitions and excessive somnolence being the domains most impacted. A non-linear dose effect was observed, whereby worse sleep-wake transitions occurred predominantly with low levels of exposure. In addition, those in the group with a light reducing pattern of PAE, compared to abstainers, experienced greater problems with sleep-wake transitions.

**Conclusion:** These findings contribute to the growing evidence that there are no safe levels of alcohol consumption during pregnancy, as even low to moderate PAE negatively impacts adolescent sleep. Identifying sleep-wake transitions and excessive somnolence as the most affected domains provides targets for both screening and intervention.

## Introduction

Healthy sleep is essential for child and adolescent development, with a vast literature highlighting the importance of sleep for neurological, physiological, cognitive, behavioural, and emotional development (see ^1^ and ^2^ for reviews). Poor sleep, which includes sleep that is insufficient, inconsistent, not restorative, disrupted, or poorly timed ^3^, has been associated with disordered development in each of these domains. For example, poor sleep has been linked to deficits in sustained attention and learning ^4,5^, decreased immune system functioning ^6,7^, externalising and internalising problems ^8^, and poor academic performance ^9^.

Many factors can negatively impact sleep in children and adolescents, one of which is prenatal alcohol exposure (PAE). Globally, 9.8% of women consume alcohol during pregnancy, although this prevalence varies significantly by country and region ^10^. Of relevance to the present study, the estimated prevalence of alcohol consumption during pregnancy in the United States is 14.8% ^10^. When a pregnant woman drinks alcohol, it passes through the placenta into the circulation of the developing fetus, and the blood alcohol concentration (BAC) in the mother and fetus quickly equilibrate. Although alcohol is easily metabolized by adults, the fetus lacks the necessary enzymes. Consequently, the alcohol is only metabolised after circulating back into the maternal blood supply ^11^, resulting in a BAC in the fetus that can be even higher than that in the mother and can remain so for a prolonged period ^12^,^13^. Alcohol is teratogenic, affecting the growth and development, and later function, of the brain and other organs. One of the most disabling potential consequences of PAE is Fetal Alcohol Spectrum Disorder (FASD; ^14^. FASD is a lifelong disorder associated with aberrations in development and function of the brain and central nervous system, manifesting as problems in psychological, cognitive, motor, and behavioural function ^15–17^. Congenital anomalies and growth restriction may also occur ^14^.

Sleep problems are a commonly reported concern for children and adolescents with FASD ^18^, with some studies indicating that as many as 58%-85% of those with FASD (aged 4-18 years) experience sleep disturbances ^19,20^. These rates are higher than has been documented in typically developing children (e.g., up to 40% ^21,22^). The type of sleep problems experienced vary, the most common being difficulties falling asleep and staying asleep ^23–27^. Other problems such as low sleep efficiency ^23^, parasomnias ^19,25^ and sleep disordered breathing ^25^, are also reported. Moreover, these sleep difficulties negatively impact the individual’s daytime functioning, further exacerbating their cognitive, psychological, and behavioural impairments ^28^, and are strongly associated with poor caregiver well-being and family quality of life ^26^.

Not all offspring exposed to alcohol prenatally will be born with FASD. Estimates suggest that one in every thirteen women who consume alcohol while pregnant will give birth to a child with FASD ^29^. Heavy PAE, particularly binge drinking, and PAE during periods of rapid fetal brain development pose the greatest risk ^30,31^. Results of several studies examining the impact of heavy PAE on sleep in children without a diagnosis of FASD align with those in children with a diagnosis of FASD. In a population-based sample of infants, Alvik et al. ^32^ found that binge drinking at least once a week during the first 6 weeks of pregnancy strongly predicted sleep problems, conceptualized as short sleep durations and sleep interruptions, during infancy. In addition, alcohol use during pregnancy at >1 drink per day has been linked to increased rates of nightmares and sleep talking ^33^, night wakings, parasomnias ^34^, and overall sleep problems ^35^ in childhood. A notable exception is a longitudinal study by Stone et al. ^36^ which found no effect of PAE on sleep outcomes in exposed children aged 1-12 years of age.

Although there is an established literature on the associations between heavy PAE and FASD on offspring outcomes, less is known about the impact of low and moderate levels of alcohol consumption during pregnancy. Recently, Lees et al. ^37^ undertook one of the most comprehensive examinations of the impact of PAE on offspring outcomes in a large sample of 9–10-year-old children, where those with PAE were exposed to, on average, 26.9 (SD = 24.5) drinks across gestation (i.e., <1 drink per week). They found that PAE was associated with altered brain structure, and more psychological and behavioural problems, compared to those without PAE. This highlights the importance of examining the whole spectrum of PAE. To our knowledge, only one study has examined the impact of low and moderate, alongside heavy, PAE on offspring sleep outcomes. This longitudinal study followed 2 year old children through to age 9, Chandler-Mather et al. ^38^ and found that heavy PAE (i.e., >7 standard drinks per week on average, or drank ≥5 drinks per occasion and on more than 2 occasions per week throughout pregnancy), but not low (i.e., <7 drinks per week on average across the entire pregnancy and never more than 1-2 drinks per occasion) or moderate (i.e., ≤ 7 standard drinks/week on average across the pregnancy and <5 drinks per occasion) PAE, was associated with more sleep problems, and that these problems persisted across childhood. Notably these studies focus on childhood, and no studies have explored the impact of low and moderate PAE on offspring sleep outcomes during adolescence, a critical developmental period with unique environmental (i.e. school demands) and biological (i.e. changes to circadian processes) changes that impact sleep ^39^. As such, further research is required to advance our understanding of how low and moderate PAE might impact adolescent sleep outcomes.

In the present study we aim to examine the long-term impacts of alcohol use on sleep outcomes in a large US sample (N=10,336) of early adolescents (12-13-year-olds). PAE will be conceptualised in three ways, including the presence or absence of PAE, total drinks consumed during pregnancy (i.e., dose), and patterns of PAE (abstinence, light drinking before knowing of pregnancy, light drinking throughout pregnancy, heavy drinking before knowing of pregnancy). Sleep will be assessed using a validated assessment tool that measures overall sleep problems and six subdomains (disorders of initiating and maintaining sleep, sleep breathing disorders, disorders of arousal, sleep-wake transition disorders, disorders of excessive somnolence, and sleep hyperhidrosis). The use of the Adolescent Brain Cognitive Development (ABCD) study means we have access to a large cohort with extensive measures, allowing us to comprehensively assess PAE and sleep outcomes as well as control for potentially confounding factors, including birth related, environmental, and medical factors. Taken together, this will allow for a rigorous examination of associations between PAE and sleep.

We tested three pre-registered hypotheses (https://osf.io/sncw7/metadata). First, given the established associations between PAE and sleep problems, we hypothesised that adolescents who were exposed to alcohol at any time during pregnancy would report higher levels of sleep problems, compared with adolescents without PAE. Second, given that heavy PAE is typically associated with worse sleep outcomes, we hypothesised that higher doses of PAE would be associated with more sleep problems during early adolescence. Third, given prior evidence suggesting that the pattern of PAE is important in terms of outcomes, we hypothesised that different patterns of PAE would be differentially associated with offspring sleep disorders.

## Methods

### Participants

The ABCD Study is a multi-site (N = 21) longitudinal study tracking the biological and behavioural development of children from the ages of 9-10 through adolescence and into adulthood. For this study, we used data from the ABCD data release 5.1. PAE variables and covariates were derived from the baseline assessment wave (ages 9-10), while the sleep variables were obtained from the fourth assessment wave (ages 12-13), at which data was collected from 10,336 participants. We also used the fourth assessment wave to fill a critical gap in the literature, namely investigating the impact of PAE, including low and moderate levels, on adolescent sleep outcomes. Parents/caregivers provided signed informed consent and all participants gave assent. The ABCD protocol was approved by the centralised institutional review board (IRB) at the University of California, San Diego and by the IRBs at the 21 sites.

### Measures

#### Sleep

Sleep outcomes were measured using the parent-reported Sleep Disturbance Scale for Children (SDSC ^40^). The SDSC is a 26-item questionnaire designed to assess multiple dimensions of childhood sleep, including: i) disorders of initiating and maintaining sleep, ii) sleep breathing disorders, iii) disorders of arousal, iv) sleep-wake transition disorders, v) disorders of excessive somnolence, and vi) sleep hyperhidrosis (sweating). Parents respond on a 5-point Likert scale (1=Never, 2=Occasionally, 3=Sometimes, 4=Often, 5=Always), about the degree to which their child experiences each of these sleep disturbances. A summed SDSC score was also calculated to reflect overall levels of sleep disturbance. For all SDSC measures and higher scores reflect greater sleep disturbances. The SDSC performed well in a review of all childhood sleep scales ^41^ and has been shown to have good internal consistency (α = 0.71 to 0.79) and test-retest reliability (r = 0.71) ^40,42,43^. In the present study, Cronbach’s α values, a measure of internal consistency, was 0.83 for overall sleep, and ranged from 0.36 to 0.81 for the sleep subscales.

#### Prenatal alcohol exposure

PAE was measured using a modified version of the Developmental History Questionnaire (DHQ ^44,45^). Mothers were asked to retrospectively report on their/the child’s biological mothers’ alcohol use before and after knowledge of pregnancy (yes/no), the maximum number of drinks consumed on a single occasion before and after knowledge of pregnancy, and the average number of drinks consumed per week during pregnancy before and after knowledge of pregnancy.

Using these questions on PAE, we calculated the following three variables according to methods outlined by Lees et al. ^37^ and used them as predictor variables in subsequent analyses: 1) a binary categorical variable to reflect any alcohol use at any time during pregnancy; 2) an estimate of the total number of drinks consumed during pregnancy which was winsorised at 1.5% to convert outliers, as was done in Lees et al. (2020); and 3) categories of common exposure patterns that have been previously applied to the ABCD data ^37^ and are based on an established classification of PAE ^46^. These categories include abstainers (abstinent throughout pregnancy), light reducers (light alcohol consumption before knowledge of pregnancy and abstinent after knowing of pregnancy), light stable users (light alcohol consumption throughout pregnancy), and heavier reducers (moderate, heavy or binge drinking before knowledge of pregnancy and abstinent or light drinking after knowing of pregnancy). Further detail on the calculation of these variables is provided in the supplementary materials.

#### Fixed and random effect covariates

We adjusted for both fixed and random effect covariates. Included fixed covariates previously identified by Lees et al. ^37^ were: birth weight (in ounces); premature status (yes/no); week of maternal pregnancy knowledge; maternal age at time of birth (in years); maternal use of other substances during pregnancy (yes/no) with tobacco, cannabis, cocaine, and heroin each being included as separate variables; history of maternal depression (yes/no); sex of adolescent at birth (female/male); adolescent race/ethnicity (White, Black, Hispanic, Asian, Other); adolescent age at time of assessment (in years); maternal history of depression (yes/no); highest level of parental education (<high school diploma, high school diploma or equivalent, college, bachelor’s degree, postgraduate degree). We also included the following fixed covariates that have been identified in the sleep literature as having evidence of a prior association with sleep outcomes: adolescent asthma (yes/no); adolescent brain injury (yes/no); adolescent cerebral palsy (yes/no); obesity (yes/no); history of paternal depression (yes/no); history of maternal and paternal anxiety (yes/no); adverse life events (total number of stressful life events perceived by the adolescent to be bad assessed using the Adverse Life Events Scale); family conflict (assessed using Family Conflict subscale of the Family Environment Scale ^47^); and caregiver acceptance (Child Report of Behaviour Inventory ^48^). The ABCD data includes 8,546 families and 21 data collection sites (ABCD originally recruited across 22 sites. However, one site was dropped due to recruitment issues, leaving 21 current sites). To account for the clustered nature of the data, random effects for family and site were included.

### Statistical Analyses

#### Missing data

Missing data in the independent variables and covariates were addressed using multiple imputation. Specifically, we used the *mice* package ^49^ in R v4.4.1 ^50^. to derive multiply imputed datasets. In line with the rule of thumb whereby the number of imputations should be at least equal to the highest percent of missing data ^51–54^, which in this study was 11.06%, we derived 11 imputations. Missing data for the outcome variable was addressed using full information maximum likelihood (FIML) in the statistical models.

#### Statistical analysis

To answer our research questions, a series of 21 models were run in R v4.4.1 ^50^. For the analyses focusing on the binomial (binary PAE; hypothesis 1) and multinomial PAE variables (PAE patterns; hypothesis 3), generalised linear mixed models were conducted using the multiply imputed data and the *lme4* package ^55^ in R. Final estimates were obtained through aggregation procedures that follow Rubin’s rules using the *parameters* ^56^ and *miceadds* package ^57^ in R. For the analyses focusing on the continuous (total drinks; hypothesis 2) PAE variable to investigate dose-dependent associations, we conducted a series of generalised additive mixed models (GAMMs) using the *mgcv* package ^58^ in R. The observed data was used for the GAMMs as they produce F statistics and effective degrees of freedom (and their corresponding p-values) which are not suitable for pooling using Rubin’s rules following multiple imputation^59^. For all predictors, separate models were run for the seven sleep outcomes and each model included the covariates and random effects listed above. The multinomial analyses also included the winsorised total drinks variable as a covariate, to get a more specific estimation of the effect of PAE timing, rather than volume, on sleep outcomes. To account for multiplicity, the False Discovery Rate (FDR ^60^) was used to correct for the 7 models with binary PAE, 7 models with continuous PAE and 21 models with PAE patterns, and the FDR adjusted p-values are reported. Statistical significance was set at *p*_*(FDR)*_ < 0.05. The syntax is available on GitHub (https://github.com/emmakdevine/ABCD_Sleep_Paper).

### Sensitivity Analyses

#### E-values

For each statistically significant outcome observed from the generalised linear mixed models (those focusing on the binary PAE and PAE patterns variables), we calculated E-values for the estimate and for the limit of the confidence interval closest to the null to assess the sensitivity of the results to potential unmeasured confounding ^60^. Larger E-values were interpreted to mean that considerable unmeasured confounding would be needed to explain away an effect estimate. In contrast, a small E-value indicates that a small amount of confounding would be needed to explain away an effect estimate, noting that what constitutes a large E-value is context dependent ^61^.

#### Inverse probability weighting

Inverse probability weighting (IPW) ^62^ was used as a sensitivity analysis to further investigate the effect of PAE exposure (yes/no) on our outcomes of interest. IPW is an alternative to covariate adjustment in attempting to control for the effects of confounders and thus deliver causal insights ^63^. Specifically, it re-weights individuals to attempt to balance characteristics between the exposed and non-exposed groups ^64^. First, we fit a logistic regression model, regressing our binary PAE variable on all covariates included in the main analysis, from which we obtained propensity scores.

Inverse probability weights were calculated from the propensity scores. Standardised mean differences between groups for were calculated for all covariates before and after weighting to assess how well the groups are balanced (a standardised difference <10% is considered a negligible imbalance between groups). Kolmogorov-Smirnov statistics were used to ascertain that the distribution and variance were similar across groups (a Kolmogorov-Smirnov statistic <0.05 indicating better balance ^64^). The main analyses for binary PAE (described above in the *Statistical Analysis* section) were then repeated including probability weights and adjusting for covariates (i.e., a doubly robust analysis), with robust standard errors calculated to account for the uncertainty added by the propensity scores. The results were compared to those from the unweighted models. The R packages *WeightIt* ^65^ and *MatchThem* ^66^ were used for the analyses.

#### Deviations from the pre-registration

The analysis presented here differed in four ways from the pre-registered protocol. First, we used the ABCD 5.1 data release instead of the originally proposed 4.0 release, as a new wave of data became available after pre-registration. This changed participants’ age ranges from 11-12 to 12-13 years, which is better suited to investigating the impact of PAE on adolescent sleep outcomes. Second, paternal depression was included as a covariate in all models, given established links between paternal depression and offspring sleep (e.g., ^67,68^. Third, the total drinks variable used to assess a dose-response effect was included as an additional covariate in the models assessing patterns of PAE exposure (hypothesis 3), to more accurately reflect the impact of patterns of PAE specifically on sleep outcomes. Fourth, due to computational constraints on bootstrapping multiply imputed data, we used robust standard errors to calculate the confidence intervals for the IPW sensitivity analyses.

## Results

### Study sample

The summary statistics for the predictor and outcome variables included in this study, prior to multiple imputation, are presented in Table 1 (see Table S2 in the supplementary materials for the summary statistics for all variables included in the study). Of the 10,336 adolescents included in this study (M_age_ = 12.91, SD_age_ = 0.65, 47.5% female at birth), 25.5% (N = 2,582) had parent-reported prenatal exposure to alcohol. The winsorised estimated total number of drinks consumed during pregnancy ranged from 0 to 116 and among those who had consumed alcohol, the mean number of drinks consumed throughout pregnancy was 26.47 (SD=28.18). Regarding the common exposure patterns, most participants were classified in the abstinent group (N=7175), followed by light reducers (N=1278), and heavy reducers (N = 787) and the light stable group with the smallest number of participants (N = 94).

### Binary prenatal alcohol exposure associations

Results of the covariate adjusted linear mixed models using the multiply imputed data are provided in Table 2. Adolescents who were prenatally exposed to any alcohol experienced greater overall sleep disturbances (β = 0.517, 95% CI = 0.127 – 0.906, *p*_*FDR*_ = 0.021), compared to unexposed adolescents. Using the sleep disturbance subscales, adolescents exposed to any alcohol exhibited more problems on the sleep wake transition disorder scale (β = 0.210, 95% CI = 0.101 – 0.318, *p*_*FDR*_ = 0.001) and on the disorders of excessive somnolence scale (β = 0.200, 95% CI = 0.069 – 0.331, *p*_*FDR*_ = 0.011), compared to unexposed adolescents. There were no differences between the groups on the remaining sleep subscales (i.e., disorders of initiating and maintaining sleep, sleep breathing disorder, disorders of arousal, or sleep hyperhidrosis).

### Dose-dependent associations

Table 3 presents the results from the 7 GAMM models investigating the associations between PAE measured by the total number of drinks consumed during pregnancy (total drinks) and sleep outcomes. A statistically significant non-linear association was observed between total drinks and sleep-wake transition disorders (EDF = 5.351, F = 2.886, *p*_*FDR*_ = 0.049). By inspecting the plot of this association (see Figure 1) we see that an increase in symptoms of sleep-wake transition disorder occurs predominantly with low level alcohol exposure (0 to 15 drinks). No dose dependent effects of PAE on overall disordered sleep, disorders of initiating and maintaining sleep, sleep breathing disorders, disorders of arousal, disorders of excessive somnolence, or sleep hyperhidrosis, were observed.

### PAE exposure pattern associations

Looking at the PAE exposure patterns, on average, women in the *light stable* group consumed 48.30 (SD = 31.23) drinks throughout pregnancy, those in the *light reducing* group consumed 15.65 (SD = 16.54) drinks, and those in the *heavy reducing* consumed 39.66 (SD = 32.52) drinks.

Results from linear mixed models adjusting for both covariates and random effects (see Table 4) showed that, compared to unexposed adolescents, the *light reducing* and *heavy reducing* PAE groups experienced worse overall sleep outcomes, however after FDR correction only the effect for the *light reducing* group remained statistically significant (β = 0.801, 95% CI = 0.285 - 1.317, *p*_*FDR*_ = 0.021). Regarding the sleep disorders subscales, *light reducing* and *heavy reducing* groups experienced greater problems with sleep wake transition disorders, compared to those in the abstinent group. However, like with the overall sleep measure, only the effect for the *light reducing* group remained statistically significant after FDR correction (β = 0.296, 95% CI = 0.152 – 0.439, *p*_*FDR*_ = 0.002). All PAE groups experienced greater problems with disorders of excessive somnolence, compared to those in the abstinent group, however none survived FDR correction. No other differences were observed between exposure groups and unexposed adolescents on the sleep outcomes.

### Sensitivity analyses

#### E-values

The E-values for the statistically significant associations between binary PAE and sleep outcomes, and the patterns of PAE groups and sleep outcomes are reported in Table 5. E-values ranged from 1.690 to 3.546, indicating that to fully account for the observed association, one or more unmeasured confounders would need to considerably increase the probability of an individual being prenatally exposed to alcohol and the probability of an individual experiencing higher overall disordered sleep, above and beyond the measured covariates, to account for the observed associations. Consider, for example, the association between the light reducer PAE group and the total summary score for all sleep disorders, which has an E-value of 3.546. This suggests that one or more unmeasured confounders would need to more than triple both the probability of an individual being exposed to alcohol prenatally and the probability that they experience greater levels of sleep disorders to fully explain this association. Similarly, the E-value of 1.715 for the association between binary PAE and sleep-wake transition disordered sleep indicates that an unmeasured confounder would need to almost double the probability of an individual having PAE and the probability that they experience greater sleep-wake transition disordered sleep to discount this association. Given the inclusion of several robustly measured covariates in the analyses, we anticipate the existence of unmeasured confounders of this magnitude to be unlikely, thus adding to the robustness of these findings.

#### Inverse probability weighting

The IPW adjustment was effective in emulating a more balanced sample, as indicated by the improved covariate balance shown in Figure S1 in the supplementary materials. Results from the doubly robust models (see Table 6) were similar to those from the unweighted models, which indicates that the observed associations are not driven by differences in measured characteristics between groups. Specifically, adolescents with PAE experienced greater overall sleep disturbances (β = 0.770, 95% CI = 0.260 – 1.281, *p*_*FDR*_ = 0.007) compared to unexposed adolescents. Using the sleep disturbance subscales, alcohol exposed adolescents exhibited greater problems on the sleep wake transition disorder scale (β = 0.274, 95% CI = 0.134 – 0.415, *p*_*FDR*_ = <0.001) and on disorders of excessive somnolence (β = 0.249, 95% CI = 0.099 – 0.400, *p*_*FDR*_ = 0.004), compared to unexposed adolescents. There were no differences between the groups on the remaining sleep subscales (i.e., disorders of initiating and maintaining sleep, sleep breathing disorder, disorders of arousal, or sleep hyperhidrosis).

## Discussion

There is an established literature linking both FASD and heavy PAE with poor sleep outcomes, particularly among infant and child populations (e.g., ^19,24,32,34^). However, less is known about how lower levels of PAE might also impact sleep outcomes, despite a growing consensus that even low levels of PAE can cause significant offspring harms. Moreover, it is unclear how PAE is associated with sleep outcomes in adolescent populations, despite the critical importance of sleep on adolescent development ^1,2^. As such, this paper sought to address these gaps by investigating the associations between PAE, including low levels of PAE, and sleep in a large sample of adolescents.

As hypothesised, adolescents with PAE, compared to those without, experienced worse sleep outcomes. While some studies in populations with FASD (e.g., ^24^) have identified associated across all sleep domains, our findings in this sample of primarily low to moderate PAE (26.47 drinks consumed across gestation) revealed the impact to be more localised, with sleep-wake transitions and excessive somnolence emerging as the domains most impacted by PAE. The robustness of these findings across rigorous sensitivity analyses underscores the reliability of these associations, highlighting that even low levels of PAE can have targeted and meaningful effects on sleep outcomes in adolescence. While the effect sizes for significant findings are quite small, it is important to note that this is not uncommon in ABCD data ^69,70^ and that these small effects are still relevant at a public health level, especially given the high prevalence of alcohol consumption during pregnancy in the population ^10^.

Sleep-wake transition disorders encompass any unusual or undesirable physical event or experience that occurs during the transition between sleep and wake states (e.g., sleep-talking, twitching or jerking movements, teeth grinding, and vivid dreams). They tend to impact the quality of an individual’s sleep, more so than the quantity, and people who experience problems with sleep-wake transitions often report fragmented, insufficient, and poor-quality sleep ^71^ and reduction in the restorative nature of their sleep ^71,72^. The negative impacts of sleep-wake transition disorders on sleep quality may underlie our finding that PAE is also associated with disorders of excessive somnolence, defined as an inability to stay awake, particularly during the day ^73^. It is widely accepted that PAE alters an individual’s neurobiology (e.g., ^37,74^), which subsequently impacts sleep. The field would benefit from further research investigating which brain regions or processes mediate the associations between PAE and problems with sleep-wake transitions and excessive somnolence specifically.

This study also sought to investigate the impact of dose and exposure pattern of PAE on sleep outcomes. Specifically, we hypothesised that a higher dose of PAE would lead to worse offspring sleep outcomes, however this hypothesis was not supported. Indeed, results showed that lower levels of PAE have the greatest impact, particularly on sleep-wake transition disorders. This conflicts with existing research showing stronger associations between heavy PAE, rather than low-moderate PAE, and sleep outcomes (e.g., ^32^-34,^38^). However, our findings align with research showing that even low level PAE can incur risk (e.g., ^37^). Similarly, in investigating the associations between patterns of PAE and sleep outcomes, our results showed that adolescents in the light reducing PAE group (i.e., those whose mothers drank low levels of alcohol before pregnancy knowledge and were abstinent after knowledge of pregnancy) experienced worse sleep outcomes, specifically sleep-wake transition disordered sleep. Notably, adolescents in the light stable and heavy reducing PAE groups did not experience worse sleep outcomes. These findings may reflect unique circumstances or characteristics in the light reducing group that may be driving these findings. For example, this group of mothers may have reduced their alcohol consumption during pregnancy due to underlying health concerns of complications, which may reflect a more complex health profile that could influence fetal development, and consequently offspring sleep patterns.

There are several strengths and limitations of the current study which should be considered when interpreting these findings. Strengths include the use of the ABCD data which constitutes a large, representative, and well-characterised sample. The ABCD data also allowed us to conceptualise PAE in multiple ways, allowing for a nuanced examination of the impact of PAE on sleep outcomes. The present study also used sophisticated and robust analysis methods, including causal methods such as IPW. Regarding the limitations, although commonly used to document PAE ^29^, retrospective maternal report of alcohol consumption during pregnancy may be subject to reporting bias, largely due to the stigma associated with PAE, and recall bias (see ^29^ for a review). For example, in a large Canadian study, maternal self-report of alcohol consumption during pregnancy was ten times lower than objective measures of consumption via analysis of meconium fatty acid ethyl esters ^75^. Thus, individuals categorised in the low PAE group may in fact have been exposed to substantially higher levels of PAE, which would align our findings more closely with studies that show greater adverse impact of heavier PAE on sleep. Similarly, although reliable in infant and child populations ^76,77^, parental report of sleep problems in adolescence becomes less accurate, likely due to parents having less oversight over their child’s sleep as they get older ^78^ and, as such, the reliability of the reported sleep problems may be low. Thus, the adverse impact of lower levels of PAE on sleep outcomes may reflect a genuine effect, the effect of an uncontrolled variable that impacted offspring sleep outcomes, or limitations with the data itself. Replicating these analyses in other newly established large-scale datasets, such as the Healthy Brain and Child Development (HBCD) Study ^79^, which allow for several of the above limitations to be addressed, may lend further clarity.

In addition to addressing the limitations mentioned above, future work should prioritise the development and evaluation of interventions aimed at improving sleep in populations with PAE. Sleep problems exacerbate impairments associated with PAE and potentially lead to worsening outcomes over time ^34^. Despite this, there is currently no research exploring sleep interventions for individuals with PAE, and the efficacy of existing sleep hygiene programs for this population remains unclear ^80^. Such interventions have the potential to substantially improve the quality of life of individuals with sleep problems resulting from PAE ^28^ and their families ^26^.

In conclusion, this large, rigorous study was the first to examine the impact of PAE, including low levels of PAE, on sleep outcomes in adolescence. We found that PAE is associated with worse sleep outcomes, particularly sleep-wake transitions and excessive somnolence. The results for the analyses investigating the role of PAE dose and pattern were contrary to what we expected, whereby lower levels of PAE were associated with worse sleep outcomes. We discussed how unique circumstances or characteristics in the light PAE group might be driving these findings and explored the potential strengths and limitations of the study which should be considered in interpreting the results. Further research should consider addressing these limitations, for example through objective measures of both PAE (e.g., detecting levels of PAE using baby teeth ^81,82^) and sleep (e.g., use of mobile and wearable technologies to track sleep ^83^). Efforts should be made to develop interventions with the aim of ameliorating the negative consequences of poor sleep in adolescent populations with PAE.

## Acknowledgements

This research was supported by grants from the NIH’s NIAAA (Award numbers: R01AA030575, MPI: Mewton/Squeglia; K24AA031052, PI: Squeglia). L.A.S. is supported by an NHMRC Investigator Grant Fellowship (GNT2026380)

## Data availability

This study used data from the Adolescent Brain Cognitive Development (ABCD) Study. Researchers can apply for access to the data here: https://nda.nih.gov/abcd/request-access

## References

1. Tarokh L, Saletin JM, Carskadon MA. Sleep in adolescence: Physiology, cognition and mental health. Neuroscience & Biobehavioral Reviews. 2016;70:182–188.

2. Agostini A, Centofanti S. Normal sleep in children and adolescence. Psychiatric Clinics. 2024;47(1):1–14.

3. Gariepy G, Danna S, Gobina I, et al. How are adolescents sleeping? Adolescent sleep patterns and sociodemographic differences in 24 European and North American countries. Journal of adolescent Health. 2020;66(6):S81–S88.

4. Abel T, Havekes R, Saletin JM, Walker MP. Sleep, plasticity and memory from molecules to whole-brain networks. Current biology. 2013;23(17):R774–R788.

5. Beebe DW. Cognitive, behavioral, and functional consequences of inadequate sleep in children and adolescents. Pediatric Clinics. 2011;58(3):649–665.

6. Dickstein JB, Moldofsky H. Sleep, cytokines and immune function. Sleep Medicine Reviews. 1999;3(3):219–228.

7. Orzech KM, Acebo C, Seifer R, Barker D, Carskadon MA. Sleep patterns are associated with common illness in adolescents. Journal of Sleep Research. 2014;23(2):133–142.

8. Shimizu M, Zeringue MM, Erath SA, Hinnant JB, El-Sheikh M. Trajectories of sleep problems in childhood: associations with mental health in adolescence. Sleep. 2021;44(3):zsaa190.

9. Perkinson-Gloor N, Lemola S, Grob A. Sleep duration, positive attitude toward life, and academic achievement: the role of daytime tiredness, behavioral persistence, and school start times. Journal of adolescence. 2013;36(2):311–318.

10. Popova S, Lange S, Probst C, Gmel G, Rehm J. Estimation of national, regional, and global prevalence of alcohol use during pregnancy and fetal alcohol syndrome: a systematic review and meta-analysis. The Lancet Global Health. 2017;5(3):e290–e299. doi:10.1016/s2214-109x(17)30021-9

11. Boschen K, Klintsova A. Neurotrophins in the brain: interaction with alcohol exposure during development. Vitamins and hormones. 2017;104:197–242.

12. Nava-Ocampo AA, Velázquez-Armenta Y, Brien JF, Koren G. Elimination kinetics of ethanol in pregnant women. Reproductive Toxicology. 2004;18(4):613–617.

13. van Faassen E, Niemelä O. Biochemistry of prenatal alcohol exposure. 2011;

14. Popova S, Charness ME, Burd L, et al. Fetal alcohol spectrum disorders. Nature reviews Disease primers. 2023;9(1):11.

15. Caputo C, Wood E, Jabbour L. Impact of fetal alcohol exposure on body systems: a systematic review. Birth Defects Research Part C: Embryo Today: Reviews. 2016;108(2):174–180. doi:10.1002/bdrc.21129

16. Riley EP, Infante MA, Warren KR. Fetal alcohol spectrum disorders: an overview. Neuropsychology review. 2011;21:73–80.

17. Cook JL, Green CR, Lilley CM, et al. Fetal alcohol spectrum disorder: a guideline for diagnosis across the lifespan. Cmaj. 2016;188(3):191–197.

18. Hanlon-Dearman A, Chen ML, Olson HC. Understanding and managing sleep disruption in children with fetal alcohol spectrum disorder. Biochemistry and Cell Biology. 2018;96(2):267–274. doi:10.1139/bcb-2017-0064

19. Chen ML, Olson HC, Picciano JF, Starr JR, Owens J. Sleep problems in children with fetal alcohol spectrum disorders. Journal of Clinical Sleep Medicine. 2012;8(4):421–429. doi:10.5664/jcsm.2038

20. Goril S, Zalai D, Scott L, Shapiro CM. Sleep and melatonin secretion abnormalities in children and adolescents with fetal alcohol spectrum disorders. Sleep Medicine. 2016;23:59–64. doi:10.1016/j.sleep.2016.06.002

21. Owens JA, Spirito A, McGuinn M, Nobile C. Sleep habits and sleep disturbance in elementary school-aged children. Journal of Developmental and Behavioral Pediatrics. 2000;21(1):27–36. doi:10.1097/00004703-200002000-00005

22. Gradisar M, Gardner G, Dohnt H. Recent worldwide sleep patterns and problems during adolescence: a review and meta-analysis of age, region, and sleep. Sleep medicine. 2011;12(2):110–118.

23. Mughal R, Hill CM, Joyce A, Dimitriou D. Sleep and cognition in children with fetal alcohol spectrum disorders (FASD) and children with autism spectrum disorders (ASD). Brain Sciences. 2020;10(11):863.

24. O’Rourke C, Horne RS, Nixon GM, Harris KR, Connelly A, Crichton A. Sleep Disturbance in Children with Fetal Alcohol Spectrum Disorder and the Relationship to the Neurodevelopmental Profile. Journal of Developmental & Behavioral Pediatrics. 2024;45(4):e358–e364.

25. Dylag KA, Bando B, Baran Z, et al. Sleep problems among children with Fetal Alcohol Spectrum Disorders (FASD)-an explorative study. Italian Journal of Pediatrics. 2021;47(1):113. doi:10.1186/s13052-021-01056-x

26. Hayes N, Moritz K, Reid N. Parent-reported sleep problems in school-aged children with fetal alcohol spectrum disorder: association with child behaviour, caregiver, and family functioning. Sleep Medicine. 2020;74:307–314.

27. Wengel T, Hanlon-Dearman AC, Fjeldsted B. Sleep and sensory characteristics in young children with fetal alcohol spectrum disorder. Journal of Developmental & Behavioral Pediatrics. 2011;32(5):384–392.

28. Blackmer AB, Feinstein JA. Management of sleep disorders in children with neurodevelopmental disorders: a review. Pharmacotherapy: The Journal of Human Pharmacology and Drug Therapy. 2016;36(1):84–98.

29. Lange S, Probst C, Gmel G, Rehm J, Burd L, Popova S. Global prevalence of fetal alcohol spectrum disorder among children and youth: a systematic review and meta-analysis. JAMA pediatrics. 2017;171(10):948–956.

30. May PA, Tabachnick BG, Gossage JP, et al. Maternal factors predicting cognitive and behavioral characteristics of children with fetal alcohol spectrum disorders. Journal of developmental and behavioral pediatrics. 2013;34(5):314. doi:10.1097/dbp.0b013e3182905587

31. Petrelli B, Weinberg J, Hicks GG. Effects of prenatal alcohol exposure (PAE): insights into FASD using mouse models of PAE. Biochemistry and Cell Biology. 2018;96(2):131–147.

32. Alvik A, Torgersen AM, Aalen OO, Lindemann R. Binge alcohol exposure once a week in early pregnancy predicts temperament and sleeping problems in the infant. Early Human Development. 2011;87(12):827–833. doi:10.1016/j.earlhumdev.2011.06.009

33. Shang CY, Gau SSF, Soong WT. Association between childhood sleep problems and perinatal factors, parental mental distress and behavioral problems. Journal of sleep research. 2006;15(1):63–73. doi:10.1111/j.1365-2869.2006.00492.x

34. Inkelis SM, Thomas JD. Sleep in infants and children with prenatal alcohol exposure. Alcoholism: Clinical and Experimental Research. 2018;42(8):1390–1405. doi:10.1111/acer.13803

35. Harskamp-van Ginkel MW, Kool RE, van Houtum L, et al. Potential determinants during ‘the first 1000 days of life’of sleep problems in school-aged children. Sleep Medicine. 2020;69:135–144. doi:10.1016/j.sleep.2019.12.020

36. Stone KC, LaGasse LL, Lester BM, et al. Sleep problems in children with prenatal substance exposure: the Maternal Lifestyle study. Archives of pediatrics & adolescent medicine. 2010;164(5):452–456. doi:10.1001/archpediatrics.2010.52

37. Lees B, Mewton L, Jacobus J, et al. Association of prenatal alcohol exposure with psychological, behavioral, and neurodevelopmental outcomes in children from the adolescent brain cognitive development study. American Journal of Psychiatry. 2020;177(11):1060–1072.

38. Chandler-Mather N, Occhipinti S, Donovan C, Shelton D, Dawe S. An investigation of the link between prenatal alcohol exposure and sleep problems across childhood. Drug and Alcohol Dependence. 2021;218:108412. doi:10.1016/j.drugalcdep.2020.108412

39. Crabtree VM, Williams NA. Normal sleep in children and adolescents. Child and Adolescent Psychiatric Clinics. 2009;18(4):799–811.

40. Bruni O, Ottaviano S, Guidetti V, et al. The Sleep Disturbance Scale for Children (SDSC) Construct ion and validation of an instrument to evaluate sleep disturbances in childhood and adolescence. Journal of sleep research. 1996;5(4):251–261.

41. Spruyt K, Gozal D. Pediatric sleep questionnaires as diagnostic or epidemiological tools: a review of currently available instruments. Sleep medicine reviews. 2011;15(1):19–32.

42. Ağca S, Görker I, Turan FN, Öztürk L. Validity and reliability of the Turkish version of Sleep Disturbance Scale for Children. Sleep Medicine. 2021;84:56–62.

43. Lewandowski AS, Toliver-Sokol M, Palermo TM. Evidence-based review of subjective pediatric sleep measures. Journal of pediatric psychology. 2011;36(7):780–793.

44. Kessler RC, Avenevoli S, Green J, et al. National comorbidity survey replication adolescent supplement (NCS-A): III. Concordance of DSM-IV/CIDI diagnoses with clinical reassessments. Journal of the American Academy of Child & Adolescent Psychiatry. 2009;48(4):386–399.

45. Merikangas KR, Avenevoli S, Costello EJ, Koretz D, Kessler RC. National comorbidity survey replication adolescent supplement (NCS-A): I. Background and measures. Journal of the American Academy of Child & Adolescent Psychiatry. 2009;48(4):367–379.

46. O’Leary CM, Bower C, Zubrick SR, Geelhoed E, Kurinczuk JJ, Nassar N. A new method of prenatal alcohol classification accounting for dose, pattern and timing of exposure: improving our ability to examine fetal effects from low to moderate alcohol. Journal of Epidemiology & Community Health. 2010;64(11):956–962.

47. Moos RH, Moos BS. Family environment scale manual: Development, applications, research. (No Title). 1994;

48. Schaefer ES. A configurational analysis of children’s reports of parent behavior. Journal of consulting psychology. 1965;29(6):552.

49. Van Buuren S, Groothuis-Oudshoorn K. mice: Multivariate imputation by chained equations in R. Journal of statistical software. 2011;45:1–67.

50. R: A language and environment for statistical computing.. R Foundation for Statistical Computing; 2021. https://www.R-project.org/.

51. Graham JW, Olchowski AE, Gilreath TD. How many imputations are really needed? Some practical clarifications of multiple imputation theory. Prevention science. 2007;8:206–213.

52. Bodner TE. What improves with increased missing data imputations? Structural equation modeling: a multidisciplinary journal. 2008;15(4):651–675.

53. White IR, Royston P, Wood AM. Multiple imputation using chained equations: issues and guidance for practice. Statistics in medicine. 2011;30(4):377–399.

54. Wulff JN, Jeppesen LE. Multiple imputation by chained equations in praxis: guidelines and review. Electronic Journal of Business Research Methods. 2017;15(1):41–56.

55. Bates D, Mächler M, Bolker B, Walker S. Fitting linear mixed-effects models using lme4. arXiv preprint arXiv:14065823. 2014;

56. Lüdecke D, Ben-Shachar MS, Patil I, Makowski D. Extracting, computing and exploring the parameters of statistical models using R. Journal of Open Source Software. 2020;5(53):2445.

57. miceadds: Some additional multiple imputation functions, especially for ‘mice’. Version R package version 3. 16–18. 2024. https://CRAN.R-project.org/package=miceadds

58. Wood SN. Fast stable restricted maximum likelihood and marginal likelihood estimation of semiparametric generalized linear models. Journal of the Royal Statistical Society Series B: Statistical Methodology. 2011;73(1):3–36.

59. Bolt, M. A., MaWhinney, S., Pattee, J. W., Erlandson, K. M., Badesch, D. B., & Peterson, R. A. (2022). Inference following multiple imputation for generalized additive models: an investigation of the median p-value rule with applications to the Pulmonary Hypertension Association Registry and Colorado COVID-19 hospitalization data. BMC Medical Research Methodology, 22(1), 148.

59. Benjamini Y, Hochberg Y. Controlling the false discovery rate: a practical and powerful approach to multiple testing. Journal of the Royal statistical society: series B (Methodological). 1995;57(1):289–300.

60. VanderWeele TJ, Ding P. Sensitivity analysis in observational research: introducing the E-value. Annals of internal medicine. 2017;167(4):268–274.

61. VanderWeele TJ, Ding P, Mathur M. Technical considerations in the use of the E-value. Journal of Causal Inference. 2019;7(2):20180007.

62. Rosenbaum PR, Rubin DB. The central role of the propensity score in observational studies for causal effects. Biometrika. 1983;70(1):41–55.

63. Visontay R, Squeglia LM, Sunderland M, Devine EK, Byrne H, Mewton L. Enhancing causal inference in population-based neuroimaging data in children and adolescents. Developmental Cognitive Neuroscience. 2024:101465.

64. Chesnaye NC, Stel VS, Tripepi G, et al. An introduction to inverse probability of treatment weighting in observational research. Clinical Kidney Journal. 2022;15(1):14–20.

65. WeighIt: Weighting for covariate balance in observational studies.. Version R package version 0.14.2. 2023. https://CRAN.R-project.org/package=WeightIt

66. Pishgar F, Greifer N, Leyrat C, Stuart E. MatchThem:: matching and weighting after multiple imputation. arXiv preprint arXiv:200911772. 2020;

67. Hall WA, Moynihan M, Bhagat R, Wooldridge J. Relationships between parental sleep quality, fatigue, cognitions about infant sleep, and parental depression pre and post-intervention for infant behavioral sleep problems. BMC pregnancy and childbirth. 2017;17:1–10.

68. Coles L, Thorpe K, Smith S, et al. Children’s sleep and fathers’ health and wellbeing: A systematic review. Sleep Medicine Reviews. 2022;61:101570.

69. Owens MM, Potter A, Hyatt CS, et al. Recalibrating expectations about effect size: A multi-method survey of effect sizes in the ABCD study. PloS one. 2021;16(9):e0257535.

70. Dick A, Lopez D, Watts A, et al. Meaningful associations in the adolescent brain cognitive development study. NeuroImage, 239, Article 118262. 2021.

71. Bollu PC, Kaur H. Sleep medicine: insomnia and sleep. Missouri medicine. 2019;116(1):68.

72. Medic G, Wille M, Hemels ME. Short-and long-term health consequences of sleep disruption. Nature and science of sleep. 2017:151–161.

73. Mayer G. 5 Excessive somnolence disorders. This page intentionally left blank. 2008:78.

74. Lebel C, Roussotte F, Sowell ER. Imaging the impact of prenatal alcohol exposure on the structure of the developing human brain. Neuropsychology review. 2011;21:102–118.

75. Delano K, Koren G, Zack M, Kapur BM. prevalence of Fetal Alcohol exposure by Analysis of Meconium Fatty Acid ethyl esters; A National Canadian study. Scientific reports. 2019;9(1):2298.

76. Byars KC, Yolton K, Rausch J, Lanphear B, Beebe DW. Prevalence, patterns, and persistence of sleep problems in the first 3 years of life. Pediatrics. 2012;129(2):e276–e284.

77. Sadeh A, Mindell J, Rivera L. “My child has a sleep problem”: a cross-cultural comparison of parental definitions. Sleep medicine. 2011;12(5):478–482.

78. Fatima Y, Doi SA, O’Callaghan M, Williams G, Najman JM, Mamun AA. Parent and adolescent reports in assessing adolescent sleep problems: results from a large population study. Acta paediatrica. 2016;105(9):e433–e439.

79. Volkow ND, Gordon JA, Bianchi DW, et al. The HEALthy Brain and Child Development Study (HBCD): NIH collaboration to understand the impacts of prenatal and early life experiences on brain development. Developmental Cognitive Neuroscience. 2024;69:101423.

80. Jan JE, Asante KO, Conry JL, et al. Sleep health issues for children with FASD: Clinical considerations. International journal of pediatrics. 2010;2010(1):639048.

81. Montag AC, Chambers CD, Jones KL, et al. Prenatal alcohol exposure can be determined from baby teeth: proof of concept. Birth defects research. 2022;114(14):797–804.

82. Uban KA, Horton MK, Jacobus J, et al. Biospecimens and the ABCD study: Rationale, methods of collection, measurement and early data. Developmental cognitive neuroscience. 2018;32:97–106.

83. Bagot K, Matthews SA, Mason M, et al. Current, future and potential use of mobile and wearable technologies and social media data in the ABCD study to increase understanding of contributors to child health. Developmental cognitive neuroscience. 2018;32:121–129.

