## Supplemental Files for "The impact of prenatal alcohol exposure on sleep outcomes in 10,336 young adolescents: An Adolescent Brain Cognitive Development (ABCD) Study"

**Supplementary Materials**

**Calculating the prenatal alcohol exposure (PAE) variables**:

**PAE Variables**: PAE was measured at baseline using a version of the Developmental History Questionnaire (DHQ; Kessler et al., 2009; Merikangas et al., 2009) that was modified to include questions on maternal use of substances during pregnancy. These questions included ones asking about alcohol use before and after knowledge of pregnancy, the maximum number of drinks consumed on a single occasion before and after knowledge of pregnancy, and the average number of drinks consumed per week during pregnancy before and after knowledge of pregnancy. In addition to the modified DHQ, questions were also asked to assess how far along the biological mother was when they found out they were pregnant, whether the child was born prematurely, and if so, how many weeks premature they were. These additional questions were used alongside those from the modified DHQ to calculate the 3 PAE variables used in this study. Further information on these items can be found in Table S1.

**Table S1:** Description of ABCD items used to calculate the prenatal alcohol exposure variables.

| ABCD 5.1 Table Name | Variable name | Variable label | Response options |
| --- | --- | --- | --- |
| ph_p_dhx | devhx_8_alcohol | Before you/biological mom found out you/biological mom were pregnant, but while you might have been pregnant with this child, did you use any of the following? Alcohol? | Yes/no/don’t know |
| ph_p_dhx | devhx_9_alcohol | Once you/biological mom knew you/she were pregnant, were you/biological mom using any of the following? Alcohol? | Yes/no/don’t know |
| ph_p_dhx | devhx_8_alchohol_avg | Before knowledge of pregnancy what was the average number of drinks consumed per week? | Average number of drinks |
| ph_p_dhx | devhx_9_alchohol_avg | After knowing of pregnancy, what was the average number of drinks consumed per week? | Average number of drinks |
| ph_p_dhx | devhx_8_alchohol_max | Before knowing of pregnancy, what was the maximum number of drinks consumed in one sitting? | Number of drinks |
| ph_p_dhx | devhx_9_alchohol_max | After knowing of pregnancy, what was the maximum number of drinks consumed in one sitting? | Number of drinks |
| ph_p_dhx | devhx_7_p | How far along (in weeks) were you when you found out that you/biological mom were pregnant? | Number of weeks |
| ph_p_dhx | devhx_12a_p | Was the child born prematurely? | Yes/no/don’t know |
| ph_p_dhx | devhx_12_p | About how many weeks premature was the child when they were born? | 1-12, 13 = greater than 12, don’t know |

**Binary PAE**: Adolescents were categorised as having been exposed to alcohol prenatally if there was any parent-reported prenatal alcohol exposure either before or after knowledge of pregnancy. R code for the calculation of the binary PAE variable is shown below.

**Estimate of the total drinks consumed during pregnancy**: To examine the dose-dependent relationships between PAE and sleep outcomes, we calculated an estimate of the total number of drinks consumed during pregnancy following the method outlined by Lees et al. (2020). This was based on the following variables and formula, the R code for which is also provided below:

a) Average number of drinks consumed per week before pregnancy knowledge.
b) The week the mother found out they were pregnant. Two weeks were subtracted from reported week of pregnancy knowledge to adjust for conception date.
c) Average number of drinks consumed per week after pregnancy knowledge.
d) Gestational week of birth.

Estimate of total number of drinks consumed = a*b + c(d – b – 2)

**PAE exposure patterns**: According to established classifications (O’Leary et al., 2010), maternal drinking can be categorised as follows: i) abstinent (<1 standard drink/occasion throughout pregnancy); ii) light (1-2 drinks/occasions, <7 drinks/week); iii) moderate (3-4 occasions, <7drinks/week); iv) heavy (<5 drinks/occasion, 7+drinks/week); v) or binge drinking (5+ occasions) before and after knowledge of pregnancy. From these classifications, it is possible to calculate the following PAE exposure patterns (Lees et al., 2020):

1. Abstainers – abstinent throughout pregnancy
2. *Increasers – drank more after knowledge of pregnancy*
3. Light stable users – stable light alcohol consumption throughout pregnancy
4. Light reducers – light before knowing about pregnancy and abstinent after knowing of pregnancy
5. Heavier reducers:
   a. Heavier use to abstinence
   b. Heavier use to light use
6. *Heavy stable users – heavy alcohol use throughout pregnancy*

Due to small sample sizes, as was the case in Lees et al. (2020), the heavy stable users, and increasers were not included in this study.

The R syntax for the calculation of the PAE variables is available on GitHub (<https://github.com/emmakdevine/ABCD_Sleep_Paper>).

**Table S2.** Summary statistics for all variables used in the present study, using the observed data.

| Variable | N = 10,336^1^ | Unexposed Adolescents  N = 7,550^1^ | Adolescents with PAE N = 2,582^1^ | p-value^2^ |
| --- | --- | --- | --- | --- |
| **Prenatal Alcohol Exposure (PAE) variables^3^** | | | | |
| **PAE** |  |  |  |  |
| Unexposed adolescents | 7,550 (74.5%) | - | - | - |
| Exposed adolescents | 2,582 (25.5%) | - | - | - |
| Missing | 204^3^ | - | - |  |
| **Total drinks** | 6.50 (18.02) | 0.00 (0.00) | 26.47 (28.18) | <0.001 |
| Missing | 1,711 | 1,043 | 464 |  |
| **PAE patterns** |  |  |  | <0.001 |
| Abstainers | 7,175 (76.9%) | 7,172 (100.0%) | 3 (0.1%)^4^ |  |
| Light stable use | 94 (1.0%) | 0 (0.0%) | 94 (4.3%) |  |
| Light reducers | 1,278 (13.7%) | 0 (0.0%) | 1,278 (59.1%) |  |
| Heavy reducing | 787 (8.4%) | 0 (0.0%) | 787 (36.4%) |  |
| Missing | 1,002 | 378 | 420 |  |
| **Sleep variables (from Sleep Disturbance Scale for Children)** | | | | |
| **Total** | 36.01 (7.89) | 35.70 (7.83) | 36.75 (7.84) | <0.001 |
| Missing | 268 | 188 | 76 |  |
| **Disorder of initiating and maintaining sleep** | 12.23 (3.94) | 12.15 (3.94) | 12.40 (3.90) | 0.005 |
| Missing | 268 | 188 | 76 |  |
| **Sleep breathing disorder** | 3.60 (1.07) | 3.60 (1.08) | 3.60 (1.02) | 0.786 |
| Missing | 268 | 188 | 76 |  |
| **Disorder of arousal** | 3.25 (0.65) | 3.25 (0.66) | 3.27 (0.64) | 0.111 |
| Missing | 267 | 188 | 75 |  |
| **Sleep-wake transition disorder** | 7.49 (2.18) | 7.39 (2.10) | 7.76 (2.32) | <0.001 |
| Missing | 268 | 188 | 76 |  |
| **Disorder of excessive somnolence** | 7.14 (2.63) | 7.04 (2.58) | 7.41 (2.71) | <0.001 |
| Missing | 266 | 188 | 74 |  |
| **Sleep hyperhidrosis** | 2.29 (0.89) | 2.28 (0.89) | 2.31 (0.89) | 0.136 |
| Missing | 268 | 188 | 76 |  |
| **Birth related variables** | |  |  |  |
| **Premature** |  |  |  | 0.009 |
| Yes | 1,952 (19.1%) | 1,481 (19.7%) | 443 (17.3%) |  |
| **Week of pregnancy knowledge** | 6.75 (6.49) | 6.71 (6.72) | 6.83 (5.71) | 0.424 |
| Missing | 1,143 | 711 | 251 |  |
| **Birth weight (lbs)** | 7.02 (1.48) | 7.00 (1.48) | 7.11 (1.43) | 0.001 |
| Missing | 1,030 | 702 | 218 |  |
| **Mother’s age at birth** | 29.65 (6.18) | 29.51 (6.20) | 30.23 (5.92) | <0.001 |
| Missing | 206 | 105 | 19 |  |
| **Prenatal tobacco exposure** | |  |  | <0.001 |
| Yes | 1,303 (12.9%) | 638 (8.5%) | 638 (25.1%) |  |
| Missing | 245 | 40 | 39 |  |
| **Prenatal cannabis exposure** | |  |  | <0.001 |
| Yes | 549 (5.5%) | 190 (2.5%) | 347 (13.8%) |  |
| Missing | 300 | 56 | 72 |  |
| **Prenatal cocaine exposure** | |  |  | <0.001 |
| Yes | 76 (0.8%) | 9 (0.1%) | 58 (2.3%) |  |
| Missing | 247 | 18 | 61 |  |
| **Prenatal heroin exposure** | |  |  | <0.001 |
| Yes | 18 (0.2%) | 5 (0.1%) | 10 (0.4%) |  |
| Missing | 261 | 17 | 72 |  |
| **Adolescent variables** | | | | |
| **Sex at birth** |  |  |  | 0.170 |
| Female | 4,909 (47.5%) | 3,547 (47.0%) | 1,254 (48.6%) |  |
| **Age** | 12.91 (0.65) | 12.91 (0.65) | 12.91 (0.65) | 0.695 |
| Missing | 1 | 0 | 0 |  |
| **Race ethnicity** |  |  |  | <0.001 |
| White | 5,592 (54.1%) | 3,873 (51.3%) | 1,656 (64.2%) |  |
| Black | 1,361 (13.2%) | 1,089 (14.4%) | 213 (8.3%) |  |
| Hispanic | 2,075 (20.1%) | 1,635 (21.7%) | 415 (16.1%) |  |
| Asian | 221 (2.1%) | 163 (2.2%) | 28 (1.1%) |  |
| Other | 1,086 (10.5%) | 790 (10.5%) | 269 (10.4%) |  |
| Missing | 1 | 0 | 1 |  |
| **Asthma** |  |  |  | 0.190 |
| Yes | 1,780 (17.2%) | 1,318 (17.5%) | 421 (16.3%) |  |
| **Brain injury** |  |  |  | 0.524 |
| Yes | 181 (1.8%) | 127 (1.7%) | 49 (1.9%) |  |
| **Cerebral palsy** |  |  |  | >0.999 |
| Yes | 15 (0.1%) | 10 (0.1%) | 4 (0.2%) |  |
| **Obesity** |  |  |  | 0.111 |
| Yes | 516 (5.0%) | 392 (5.2%) | 113 (4.4%) |  |
| **Bad life experiences** | 2.13 (2.09) | 2.08 (2.06) | 2.27 (2.14) | <0.001 |
| **Parent variables** |  |  |  |  |
| **Parental education** |  |  |  | <0.001 |
| < High school diploma | 590 (5.7%) | 531 (7.0%) | 50 (1.9%) |  |
| High school diploma or equivalent | 989 (9.6%) | 816 (10.8%) | 158 (6.1%) |  |
| College | 2,949 (28.6%) | 2,218 (29.4%) | 684 (26.5%) |  |
| Bachelor’s degree | 3,040 (29.4%) | 2,143 (28.4%) | 843 (32.6%) |  |
| Postgraduate degree | 2,756 (26.7%) | 1,831 (24.3%) | 847 (32.8%) |  |
| Missing | 12 | 11 | 0 |  |
| **Maternal depression** |  |  |  | <0.001 |
| Yes | 2,285 (23.3%) | 1,577 (21.6%) | 656 (27.0%) |  |
| Missing | 522 | 258 | 152 |  |
| **Paternal depression** |  |  |  | <0.001 |
| Yes | 1,397 (14.6%) | 944 (13.2%) | 424 (18.1%) |  |
| Missing | 759 | 396 | 235 |  |
| **Maternal anxiety** |  |  |  | 0.002 |
| Yes | 807 (8.2%) | 561 (7.6%) | 237 (9.7%) |  |
| Missing | 466 | 200 | 135 |  |
| **Paternal anxiety** |  |  |  | 0.214 |
| Yes | 478 (4.9%) | 342 (4.7%) | 129 (5.4%) |  |
| Missing | 576 | 271 | 173 |  |
| **Family conflict** | 2.07 (1.94) | 2.07 (1.91) | 2.07 (1.99) | 0.895 |
| Missing | 15 | 11 | 3 |  |
| **Caregiver acceptance** | 2.70 (0.38) | 2.70 (0.38) | 2.70 (0.38) | 0.695 |
| Missing | 12 | 9 | 3 |  |
| ^1^n (%); Mean (SD)  ^2^Welch Two Sample t-test; Pearson's Chi-squared test  ^3^204 participants were missing data on whether they were unexposed or exposed to alcohol during pregnancy. The summary statistics presented above for the Exposed and Unexposed Groups are excluding those 204 participants, while the summary statistics for the total sample column includes these participants.  ^4^Three participants reported having consumed alcohol before knowledge of pregnancy, 0 for the maximum number of drinks consuming on a single occasion before knowledge and pregnancy and the average number of drinks consumed per week before knowledge of pregnancy, resulting in 3 participants in the PAE group meeting criteria for the abstainers PAE Group. | | | | |

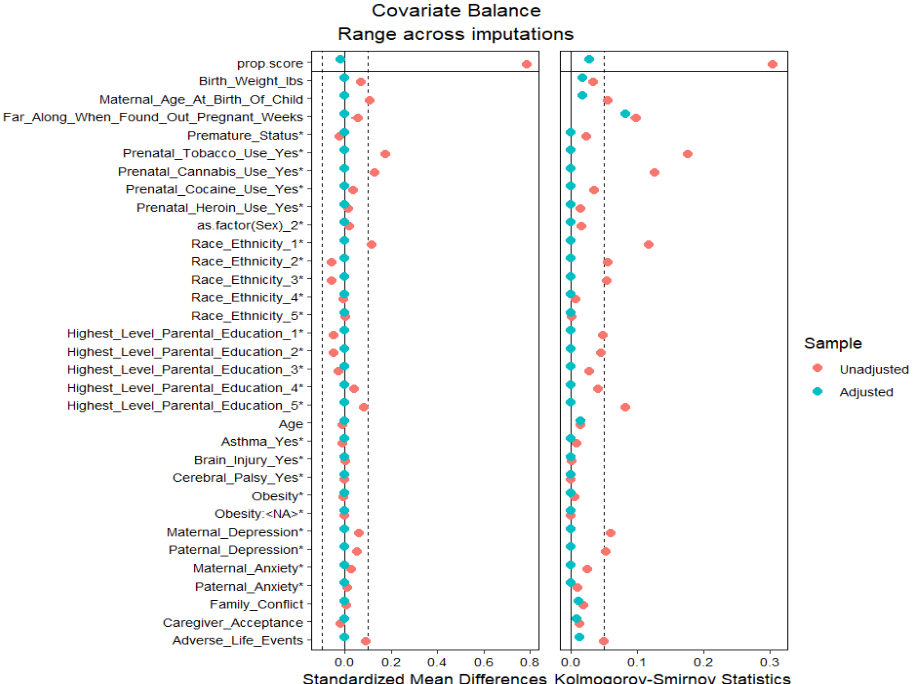

**Figure S1:** Covariate balance between PAE groups before and after weighting by the final weight. This figure displays differences in means for the two exposure groups. Variables followed by an asterisk are unstandardised categorical variables.
